# The date predicted 200.000 cases of Covid-19 in Spain using SutteARIMA

**DOI:** 10.1101/2020.05.04.20090951

**Authors:** Ansari Saleh Ahmar, Eva Boj del Val

## Abstract

The aim of this study is predicted 200.000 cases of Covid-19 in Spain. Covid-19 Spanish confirmed data obtained from Worldometer from 01 March 2020 – 17 April 2020. The data from 01 March 2020 – 10 April 2020 using to fitting with data from 11 April – 17 April 2020. For the evaluation of the forecasting accuracy measures, we use mean absolute percentage error (MAPE). Based on the results of SutteARIMA fitting data, the accuracy of SutteARIMA for the period 11 April 2020 - 17 April 2020 is 0.61% and we forecast 20.000 confirmed cases of Spain by the WHO situation report day 90/91 which is 19 April 2020 / 20 April 2020.

## 1. Introduction

Covid-19 was first reported in Wuhan, Hubei Province, China in December 2019. Covid-19 is an infectious disease caused by a new coronavirus (SARS-CoV-2) discovered in China [1]. In Spain, Covid-19 cases began to be detected on February 12, 2020. The highest addition of Covid-19 cases occurred on March 26, 2020, as many as 8271 cases and the highest daily death occurred on April 02, 2020, as many as 961 cases [2]. Based on data presented by Worldometer on April 17, 2020, the number of confirmed cases of Covid-19 in Spain was 190,839 cases or added 5891 cases from yesterday (April 16, 2020) with 20,002 total deaths and was the second highest country with confirmed cases of covid-19 in the world [2].

To see the case rate further in the future, it is necessary to forecast the data. Forecasting or predictions related to Covid-19 have been studied by various researchers: Koczkodaj, et. al predicts Covid-19 outside of China by using a simple heuristic (exponential curve) [3] and Roosa, et. Al studying about Covid-19 real-time forecast in China with generalized logistic growth model (GLM) [4]. This study forecasts 20.000 confirmed cases of Spain.

### 2. SutteARIMA

SutteARIMA is a forecasting method that combines ARIMA and α-Sutte Indicator method. The (*Z_t_*) process are an autoregressive-moving average or ARMA (p, q) model if it fulfilled [5]:

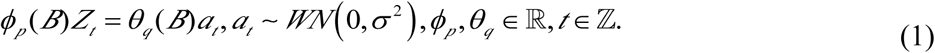

with 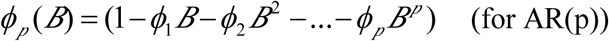
and 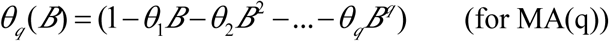

If there is a differencing then the ARIMA model becomes as follows:

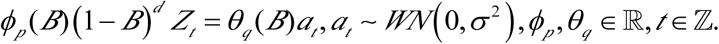

with *ϕ_p_*(*B*) = (1−*ϕ*_1_ *B− ϕ*_2_ *B*^2^ −…− *ϕ_p_B^p^*) (for AR(p)), (1 − *B)^d^* (for differencing non seasonal) and *θ_q_* (*B*) = (1 −*θ*_1_*B − θ*_2_ *B*^2^ *−*… −*θ_q_ B^q^* (for MA(q)).

The equation of the *α*-Sutte Indicator method are as follows [6,7]:

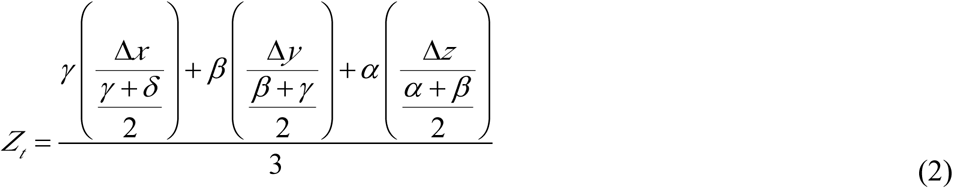

where:

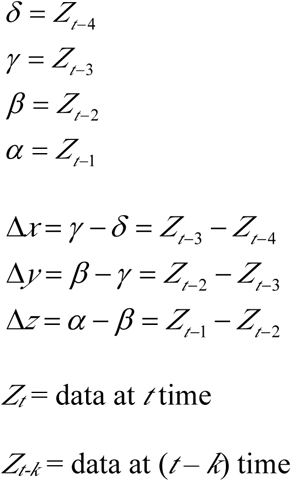

SutteARIMA is a forecasting method that combines the α-Sutte Indicator and ARIMA [8].

The equation (1), we can describe:

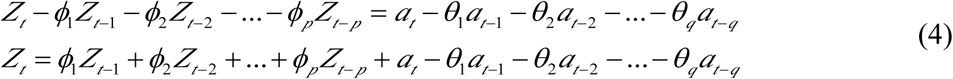

If equation (3) we reduce using backward shift operator (*B^p^Z_t_ = Z_t-p_*):

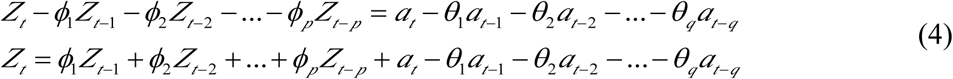

If we define:

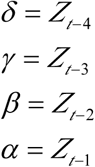

The equation (4):

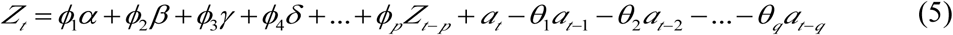

and the equation (2) we can simplify:

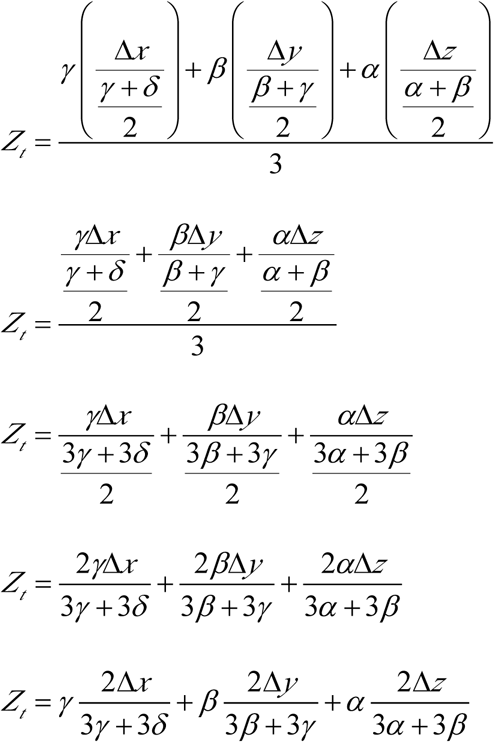

Let, Equation (2) added with Equation (5), we finding:

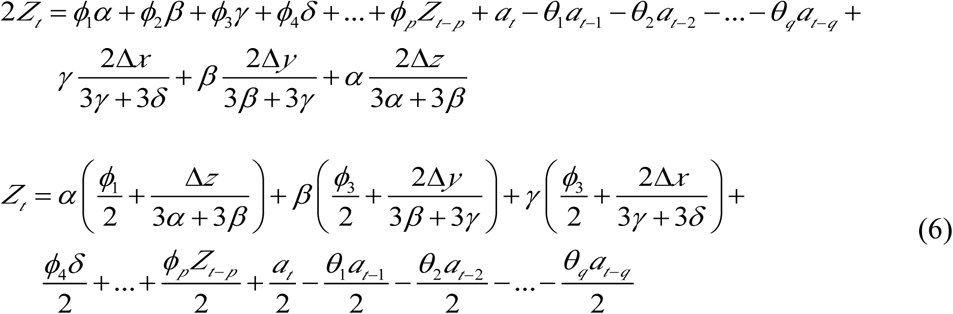

So, the equation (6) is the formula of SutteARIMA.

## 3. SutteARIMA Forecast

Covid-19 Spanish confirmed data obtained from Worldometer. Data starts from 01 March 2020 – 17 April 2020. The total confirmed cases and daily new cases in Spain can be seen in figure 1 and figure 2.

**Figure 1.**
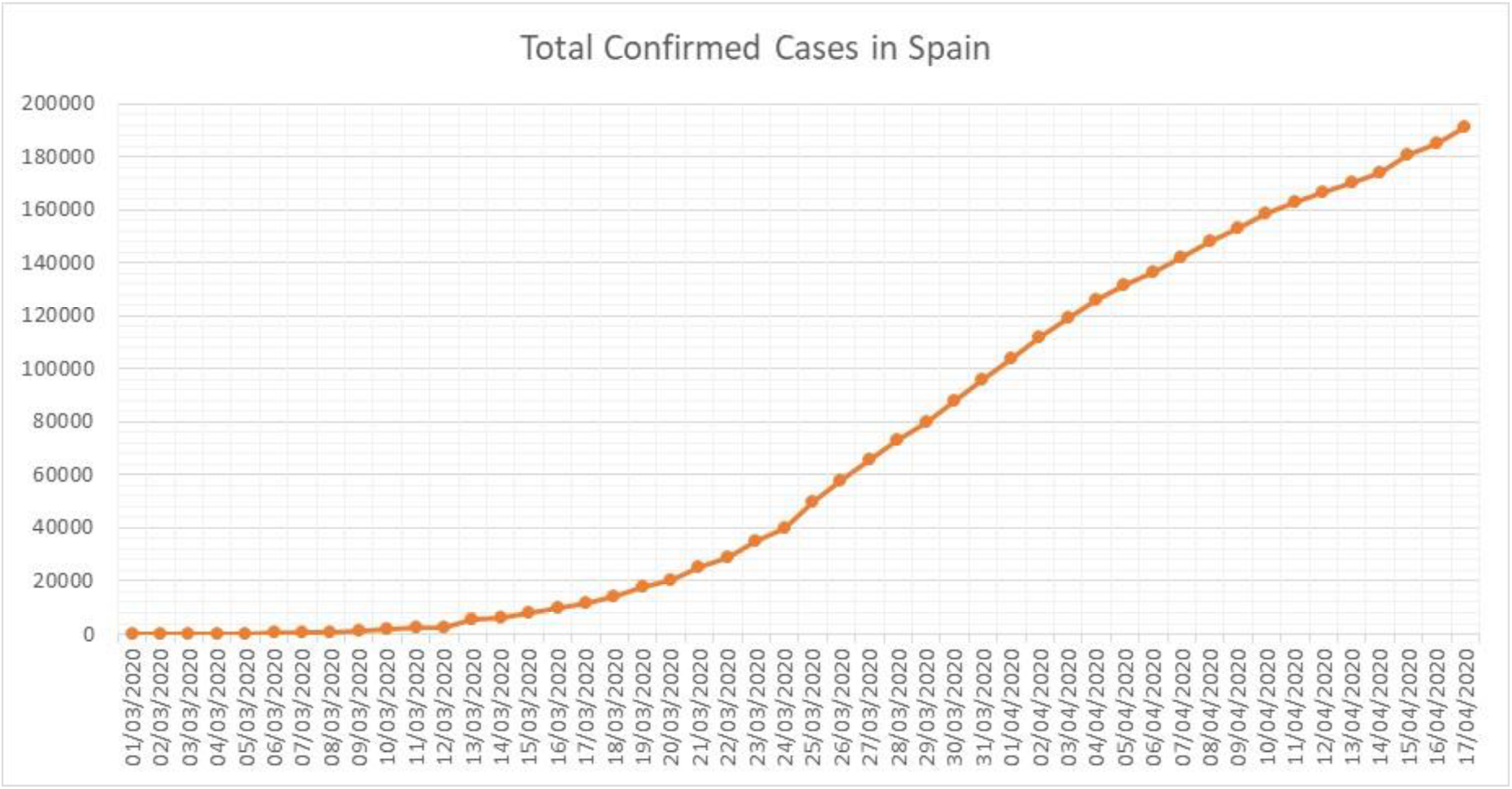
Confirmed Cases of Covid-19 in Spain (01 March 2020 – 17 April 2020)

**Figure 2.**
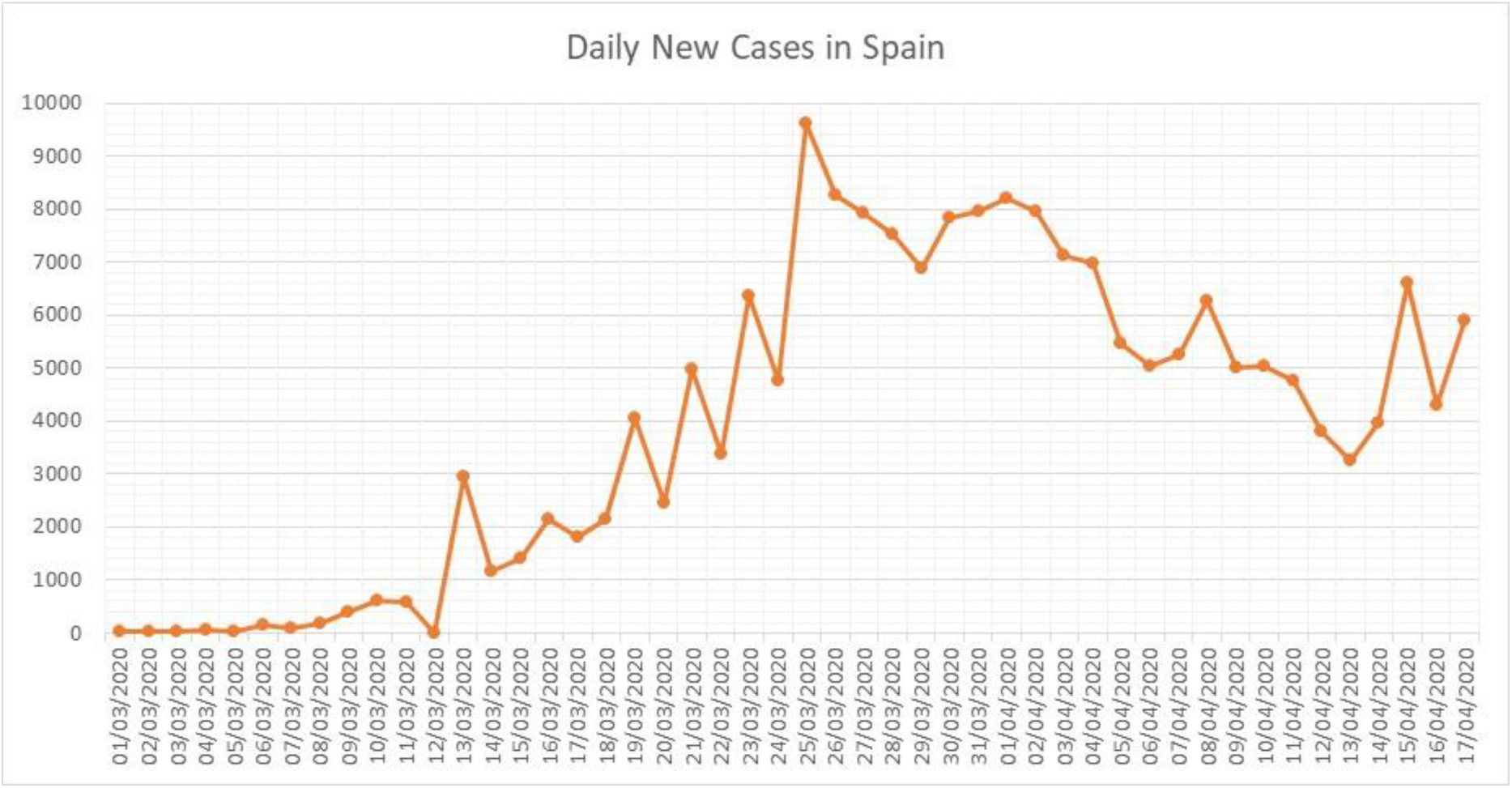
Daily New Cases of Covid-19 in Spain (01 March 2020 – 17 April 2020)

To obtain the results of forecasting, this study used the forecast and sutteForecastR of R package. To see the reliability of forecasting, first forecasting is done in the 7 previous data periods (11 April 2020 - 17 April 2020). The level of forecasting accuracy can be seen from the mean absolute percentage error (MAPE). The results of this forecast are presented in table 1.

**Table 1.** Results of Fitting Confirmed Cases of Covid-19 in Spain

| **Date** | **Actual** | **Forecast** | **APE** |
| --- | --- | --- | --- |
| **11/04/2020** | 163027 | 163817 | 0,48 |
| **12/04/2020** | 166831 | 168041 | 0,73 |
| **13/04/2020** | 170099 | 171433 | 0,78 |
| **14/04/2020** | 174060 | 174090 | 0,02 |
| **15/04/2020** | 180659 | 177778 | 1,59 |
| **16/04/2020** | 184948 | 185335 | 0,21 |
| **17/04/2020** | 190839 | 189971 | 0,45 |
| **MAPE** |  |  | **0,61** |

The table 1 shown the accuracy of SutteARIMA for the period 11 April 2020 - 17 April 2020 is 0.61%. The next step is to forecast for 20,000 cases (table 2).

**Table 2.** Forecast 20.000 Confirmed Case of Covid-19 in Spain

| **Date** | **Forecast** |
| --- | --- |
| **18/04/2020** | 196183 |
| **19/04/2020** | 201611 |
| **20/04/2020** | 207087 |

According to table 2, we predict 20.000 COVID-19 cases of Spain by the WHO situation report day 90/91 which is 19 April 2020 / 20 April 2020.

## 4. Conclusion

In particular, the SutteARIMA model is a short-term prediction model, which seems very simple and we believe that the forecast results are quite accurate with an accuracy rate of 0.5-1% for short-term predictions. We predict 20.000 COVID-19 cases of Spain by the WHO situation report day 90/91 which is 19 April 2020 / 20 April 2020.

The SutteARIMA approach that we present is based on the assumption that today’s events are influenced by the previous day using the moving average approach and the assumption that current trends can continue for the next 10 days based on the number of daily cases. The SutteARIMA results are the results of abstract mathematical forecasts and may be different in the future and the confirmed cases of COVID-19 can change in just a few days depending on whether there is an intervention occurring during the pandemic.

## Data Availability

-

## References

[1] Yang Y, Peng F, Wang R, Guan K, Jiang T, Xu G, et al. The deadly coronaviruses: The 2003 SARS pandemic and the 2020 novel coronavirus epidemic in China. J Autoimmun 2020:102434. https://doi.org/10.1016/jjaut.2020.102434.

[2] Worldometer. Spain Coronavirus 2020. https://www.worldometers.info/coronavirus/country/spain/ (accessed April 8, 2020).

[3] Koczkodaj WW, Mansournia MA, Pedrycz W, Wolny-Dominiak A, Zabrodskii PF, Strzaška D, et al. 1,000,000 cases of COVID-19 outside of China: The date predicted by a simple heuristic. Glob Epidemiol 2020:100023. https://doi.org/10.1016Zj.gloepi.2020.100023.

[4] Roosa K, Lee Y, Luo R, Kirpich A, Rothenberg R, Hyman JM, et al. Real-time forecasts of the COVID-19 epidemic in China from February 5th to February 24th, 2020. Infect Dis Model 2020;5:256–63. https://doi.org/10.1016/j.idm.2020.02.002.

[5] Wei WWS. Time Series Analysis: Univariate and Multivariate Methods. New York: Addison-Wesley Publishing Company; 1994.

[6] Ahmar AS. A Comparison of *α*-Sutte Indicator and ARIMA Methods in Renewable Energy Forecasting in Indonesia. Int J Eng Technol 2018;7:20–2.

[7] Ahmar AS, Rahman A, Mulbar U. *α*-Sutte Indicator: A new method for time series forecasting. J. Phys. Conf. Ser., vol. 1040, 2018. https://doi.org/10.1088/1742-6596/1040/1/012018.

[8] Ahmar AS, del Val EB. SutteARIMA: Short-term forecasting method, a case: Covid-19 and stock market in Spain. Sci Total Environ 2020:138883. https://doi.org/10.1016/j.scitotenv.2020.138883.

